# Polygenic susceptibility for multiple sclerosis is associated with working memory in low-performing young adults

**DOI:** 10.1101/2024.03.22.24304683

**Authors:** J. Petrovska, D. Coynel, V. Freytag, D. J.-F. de Quervain, A. Papassotiropoulos

**Affiliations:** Division of Molecular Neuroscience, Department of Biomedicine, University of Basel, CH-4055 Basel, Switzerland; Division of Cognitive Neuroscience, Department of Biomedicine, University of Basel, CH-4055 Basel, Switzerland; Research Cluster Molecular and Cognitive Neurosciences, Department of Biomedicine, University of Basel, CH-4055 Basel, Switzerland; Psychiatric University Clinics, University of Basel, CH-4055 Basel, Switzerland

**Keywords:** Multiple sclerosis (MS), genetics, polygenic risk score (PRS), working memory (WM), diffusion tensor imaging (DTI)

## Abstract

**Background:** Multiple sclerosis (MS) is a genetically complex disease with substantial heritability estimates. Besides typical clinical manifestations such as motor and sensory deficits, and fatigue, MS is characterized by structural and functional brain abnormalities, and by cognitive impairment such as decreased working memory (WM) performance.

**Objectives:** This study investigates the possible link between the polygenic risk for MS and WM performance in healthy adults aged 18 – 35 years. Additionally, it addresses in the same population the relationship between polygenic risk for MS and brain white matter properties, as represented by measures of fractional anisotropy (FA).

**Methods:** We generated a polygenic risk score (PRS) of MS and investigated its association with WM performance in a population of 3282 healthy adults, which consisted of two subsamples (*N_1_*=1803, *N_2_*=1479). The association between MS-PRS and FA was studied in the second subsample.

**Results:** MS-PRS was significantly associated with WM performance within the 10% lowest WM-performing individuals (*p* = 0.001; *p_FDR_* = 0.018). It was not significantly associated with any of the investigated FA measurements.

**Conclusions:** By identifying a genetic link between MS and WM performance, this study contributes to the understanding of the genetic complexity of MS, and hopefully to the possible identification of molecular pathways linked to cognitive deficits in MS.

## 1. Introduction

Multiple sclerosis (MS) is a complex neurodegenerative, demyelinating and inflammatory disease of the central nervous system, influenced by genetic as well as environmental factors^1^. The genetic architecture of MS is polygenic and over 200 common genetic variants have been associated with this disease in the largest genome wide association study (GWAS) to date, conducted by the International Multiple Sclerosis Genetics Consortium (IMSGC) ^2^.

Cognitive deficits are common in MS and can occur even before the onset of typical MS symptoms ^3^. MS-related deficits have been found in several cognitive domains ^4^, including working memory (WM) ^5, 6^, i.e., the capacity to temporarily maintain and manipulate a limited amount of information ^7^, and processing speed ^4^. There has been some investigation of the link between the polygenic risk for MS and cognitive functions in healthy individuals, such as nonverbal IQ in children ^8^, and a battery of several cognitive tests in adults aged 45 years or above ^9^. However, the literature on this topic is still very sparse, and none of it focuses specifically on WM. WM is a cognitive capacity with quantifiable brain functional correlates ^10^ and substantial heritability ^11^. Investigating the association between the polygenic load for MS and WM in healthy individuals may improve our understanding of the genetic architecture of this disease.

MS is also associated with structural brain changes. While the diagnostic hallmark of MS is the occurrence of demyelinating lesions, additional changes, e.g., in diffusion tensor imaging (DTI) measures of white mater properties, such as fractional anisotropy (FA) and radial diffusivity (RD) ^12^, regional grey matter changes and volume loss ^13^, have also been found in MS patients. Importantly, white matter maturation is correlated with the development of WM, and DTI-derived white matter properties have been associated with WM performance ^14, 15^.

Furthermore, the polygenic load for MS, calculated based on the latest GWAS data, has been linked to differences in global FA ^8^, as well as in spatially distinct FA clusters ^16^, in children from the general population. However, no significant associations between global or tract-specific FA differences and MS polygenic risk were found in a large sample of approximately 30’000 healthy adults over 40 years of age ^17^. These findings are consistent with previous studies on the link between the genetic risk for MS and changes in white matter properties in adults within a similar age range ^9, 18^. Some of the potential reasons for this discrepancy in findings between the studies in children and adults that have already been pointed out, e.g., impact of age-related atrophy on FA measures in the adult sample ^19^, may be partly circumvented by investigating this relationship in younger adults.

The current study is the first investigation of the link between the polygenic risk for MS, based on the largest MS GWAS currently available, and WM performance in healthy young adults. Given the relationship between WM and white matter properties, it also extends the research on the association between the polygenic load for MS and FA differences in healthy individuals, by exploring this association in young adulthood.

## 2. Materials and Methods

### 2.1. Sample

Healthy young adults (*N* = 3282) were recruited in Basel, Switzerland: Basel 1 (*N* = 1803, 67 % female; mean age: 22.56 +/- 3.67 years) and Basel 2 (*N* = 1479; 62 % female; mean age: 22.43 +/- 3.27 years). Participants did not report any neurological or psychiatric condition and did not take medication at the time of the experiment, except oral contraceptives. All participants gave written informed consent before participation and received 25 CHF/h as compensation. The ethics committee of the Cantons of Basel-Stadt and Basel-Landschaft approved the study protocols.

### 2.2. Experimental procedure and behavioural estimates

In each sample, participants performed a picture encoding task for 20 minutes. Next, they completed a verbal n-back task for ten minutes, including a 2-back and a 0-back condition. In the 0-back condition participants responded with a button press to the occurrence of the letter “x” (lower- and uppercase) in a sequence of letters (e.g., n – p – **x** …). In the 2-back condition, participants responded with a button press when the currently presented letter was identical to the letter presented two times back (e.g., n – p – **n** …). The task consisted of 12 blocks (six 0-back, six 2-back) in total. Every block consisted of 14 stimuli: three target and 11 nontarget stimuli, presented in a quasi-randomized order. Before each block started, an instruction of 5 s was shown and the block had a duration of 33 s in total. Each stimulus was presented for 500 ms with a 1500 ms interstimulus interval showing a black screen. 2-back and 0-back block sequence was randomized and there was a 20 s break after every second block. Answers were given by pressing a button to indicate each stimulus as “target” or “no target”. Of note, participants from the Basel 2 sample completed the picture encoding and the n-back task in an MRI scanner. The n-back task was followed by an unannounced free recall task in both samples. Detailed descriptions of the study procedures have been published in previous work, e.g., ^10^.

#### 2.2.1. N-back measurements

Participants’ responses with reaction time (RT) > 2000 ms, i.e., beyond the trial duration, were set to missing. In case of multiple responses for the same trial, only the first response was considered. We disregarded participants with missing responses in over 30% of all stimuli across all twelve blocks of the task, in over 30% of target stimuli in at least three blocks or in over 30% of nontarget stimuli in at least three blocks. Working memory performance was calculated as the mean accuracy and d-prime ^20^ for the 2-back condition. We also calculated mean RT for the 2-back condition and mean accuracy, d-prime and mean RT for the 0-back condition, to test for secondary effects on attention and processing speed.

### 2.3. Genotyping and genotype imputation

Genotyping data was available for both subsamples. DNA was isolated from saliva samples and processed as described in the Genome-Wide Human SNP Nsp/Sty 6.0 User Guide (Affymetrix). More information regarding the genotyping procedure can be found in previous work, e.g., ^11^. Genotype imputation was performed using the University of Michigan Imputation Server (https://imputationserver.sph.umich.edu/), independently for the Basel 1 and the Basel 2 subsample. Variants with minor allele frequency (MAF) > 0.01, call rate > 95%, imputation quality (R^2^) > 0.5, and present in both samples were considered for analysis. Subjects with genotypic data with missingness rate > 5%, excess or insufficient heterozygosity according to the majority of the sample or deviation from European ancestry background were excluded.

### 2.4. MS GWAS data

We used GWAS data from the IMSGC discovery sample meta-analysis, including 14’802 individuals with MS and 26’703 controls from 15 samples of European ancestry. Variants appearing in at least two samples were considered for analysis.

### 2.5. Polygenic risk association

Polygenic risk score (PRS) is a single value estimate of one’s genetic liability to a disease or a trait, calculated as a sum of one’s genome-wide genotypes, weighted by the corresponding genotype effect size estimates from GWAS summary statistics. For more details please see ^21^. PRSs were calculated with PRSice-2 ^22^ (v.2.3.5). The analysis was limited to the default clumping parameters setting (clump-kb 1000 kb, clump-r^2^ 0.1) and three PRSs with p-value thresholds of 5e-08, 0.05 and 1, respectively, to cover a broad range of significance. Each PRS was associated with WM performance using a quantile regression model. Unlike regular linear regression which uses the method of least squares to calculate the conditional mean of the outcome across different values of the features, quantile regression estimates the conditional median, i.e., 50^th^ percentile, of the outcome. Furthermore, quantile regression can also estimate effects at different quantiles, i.e., percentiles, including points in the upper (e.g. 90^th^ percentile) and lower (e.g. 10^th^ percentile) tails of the outcome distribution. Quantile regression is considered suitable for data with skewed distribution, and is robust to linear model assumption violations, outliers and influential points ^23^. We used a quantile regression model to calculate the association between each PRS and each behavioural estimate, respectively. Each analysis was conducted for the 50^th^ percentile, i.e. the median, as well as for the 10^th^ and 90^th^ percentile, i.e., the lowest and highest 10% of WM performers. The statistical significance threshold per analysis was set to *p* = 0.05. We used a false discovery rate (FDR) multiple-testing correction for 3 phenotypes (2-back accuracy, d prime and reaction time) * 3 PRS p-value thresholds (1, 0.05, 5 x 10-8) * 3 quantiles (0.1, 0.5, 0.9). Sex, age and the first 10 principal components (PCs) of genetic diversity were used as covariates. All variables were scaled prior to analysis. Goodness of fit was calculated for each quantile using the R^1^ measure ^24^, suitable for quantile regression, by comparing the sum of weighted deviations for each model of interest with the same sum from the respective model including only the covariates.

Given that the major histocompatibility complex (MHC) contains a substantial portion of the genetic background for MS ^2^, each PRS was calculated twice: with and without the MHC region, as done in previous work ^17^.

### 2.6. DTI

#### 2.6.1. Acquisition

Brain imaging was performed on a Siemens Magnetom Verio 3T whole-body MR unit equipped with a twelve-channel head coil. Diffusion volumes were acquired by using a single-shot echo-planar sequence, and consisted of 64 diffusion-weighted volumes (b = 900 s/mm^2^) and one unweighted volume (b = 0). Acquisition parameters are reported in the Supplementary Material.

#### 2.6.2. Pre-processing

Pre-processing of the diffusion data was based on ENIGMA’s DTI protocols (https://enigma.ini.usc.edu/protocols/dti-protocols/), using the FMRIB Software Library (FSL, v.6.0.2) and is described in the Supplementary Material. FA was the main measurement of interest, with 46 mean values per participant, following regions of interest defined in the JHU DTI-based white-matter atlas (Suppl. table 2). FA is an estimate of the directional dependence of diffusion ^25^ and represents white matter tracts properties such as fiber density and coherence within a voxel ^26^. The DTI sample consisted of 1122 participants from Basel 2.

#### 2.6.3. The ‘pothole’ method

Additionally, we applied the ‘pothole’ method ^27^ to identify non-spatially overlapping white matter abnormalities. The ‘pothole’ method was recently applied for identification of spatially distinct FA clusters related to the MS PRS in children from the general population ^16^. We used this method to identify clusters of neighbouring voxels with substantial A) FA reduction, i.e., potholes, or B) FA increase, i.e., molehills. Briefly, the FA maps normalized using the ENIGMA pipeline (see “pre-processing”) were used to derive a mean and standard deviation (SD) per voxel using all subjects and to create group mean and SD images. These images were then used to individually create a voxel-wide z-transformed image for every participant. The individual z-FA images were used to search for clusters of neighbouring voxels with z-values below a certain threshold. The z-value threshold was set to < 2 SD from the voxel-wise mean for potholes and > 2 SD from the voxel-wise mean for molehills. The cluster size was set to a minimum of 25, 50, 100, or 200 voxels, respectively, to allow testing for a broader range of MS-related white matter differences in size. The total number of potholes and molehills, respectively, were calculated as global measures of non-spatially overlapping white matter abnormalities. Furthermore, to better localize these abnormalities, the number potholes and molehills was also calculated within each white matter tract separately. For more details regarding this approach please see ^27^ and ^16^.

#### 2.6.4. Statistical analyses

The mean FA value of each region was tested for association with the MS PRS using a linear model for each region separately. Age, sex, the first 10 genetic PCs and movement during scanning (due to its negative association with brain-wide average FA, *r* = -0.06) were used as covariates. The average FA value across regions and the global and tract-based number of potholes and molehills were also checked for MS PRS association in the same manner (FDR-corrected for the number of regions tested). Of note, the PRS was calculated with a p-value threshold of 0.01 for these analyses, since this threshold was found to be optimal for identifying MS PRS - FA association in a previous study with children from the general population ^8^.

## 3. Results

### 3.1. Behavioural task performance

The distribution of participants’ scores for accuracy, d-prime and mean RT for the 2-back condition, per subsample, is presented in Figure 1. Of note, the distribution of accuracy scores was left skewed, with a larger number of participants with higher accuracy, while the distribution of mean RT was right skewed, with a larger number of participants having quicker responses. The distribution of participants’ scores for the 0-back condition, per subsample, is presented in Suppl. figure 1.

**Figure 1:**
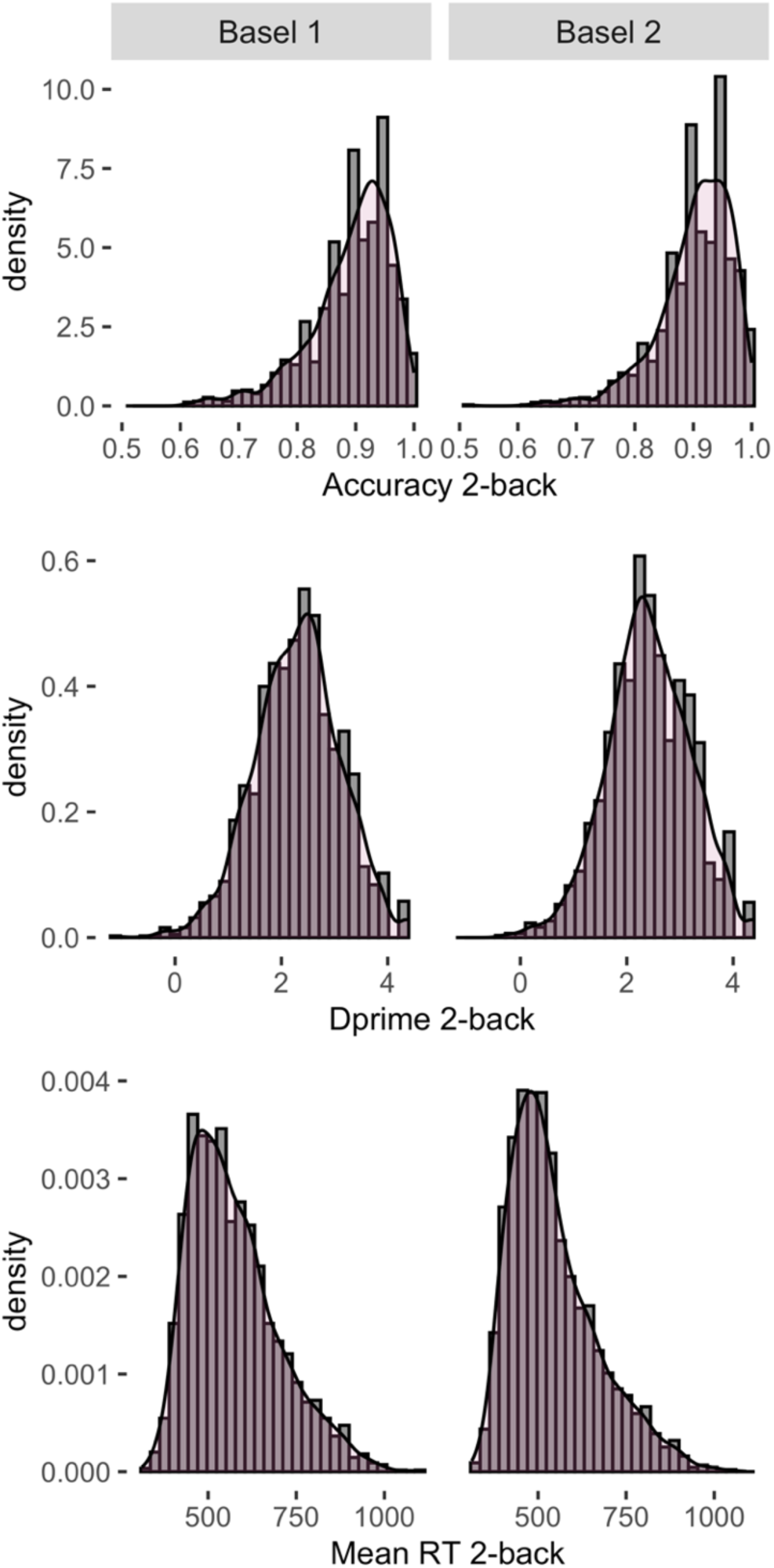
Distribution of participants’ scores for accuracy, d prime and mean reaction time (RT) of the 2-back task, per subsample.

### 3.2. MS polygenic risk association with WM

The PRS including genetic variants with nominal MS association (*p* < 0.05) had the highest association with 2-back accuracy and d-prime (Figure 2). The associations between the MS PRSs and the behavioural phenotypes from the 2-back task at the 0.1, 0.5 and 0.9 quantile, respectively, are reported in Table 1. Each association is reported for Basel 1 and Basel 2 separately, and for the entire sample, based on a common-effect meta-analysis (using the R package “meta” ^28^, v.6.2.1). The MS PRS including genetic variants with nominal MS association (*p* < 0.05) was significantly associated with WM accuracy and d-prime within the 0.1 quantile, i.e., the lowest 10% of performers, in the entire sample, after an FDR multiple testing correction, accuracy: *p* = 0.001; *p_FDR_* = 0.018; d-prime: *p* = 0.001; *p_FDR_* = 0.018, and nominally significant (*p* < 0.05) in Basel 1 and Basel 2, separately, accuracy: *p_Basel 1_* = 0.046; *p_Basel 2_* = 0.011; d-prime: *p_Basel 1_* = 0.012; *p_Basel 2_* = 0.026 (Figure 2 and Figure 3). Of note, these associations remained significant when excluding the MHC region from the PRS (Suppl. table 2). There were no other significant MS PRS associations after multiple-testing correction for 2-back accuracy, d-prime and mean RT (Table 1), or for 0-back accuracy, d-prime and mean RT (Suppl. table 3 and Suppl. table 4), in the entire sample.

**Figure 2:**
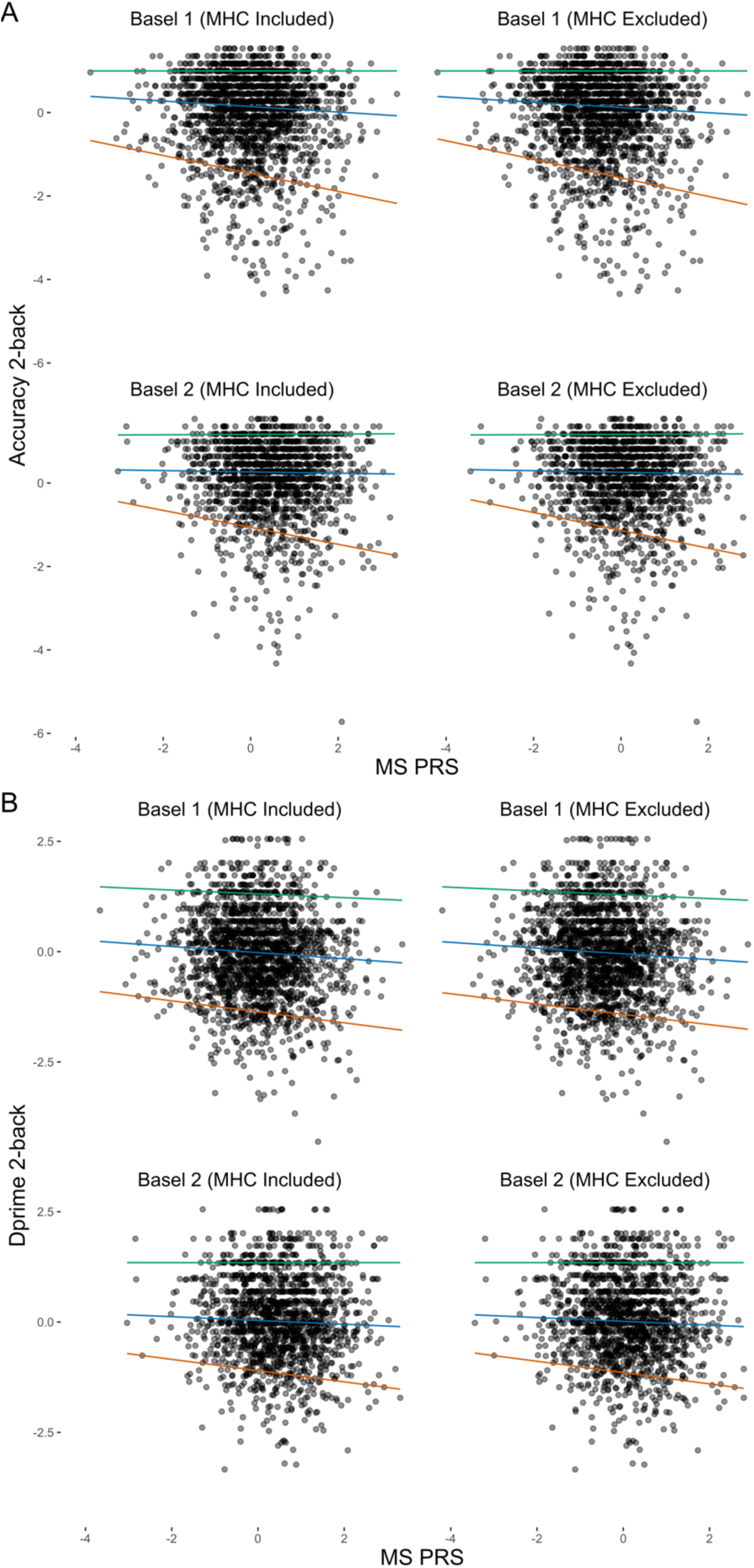
The association between the MS PRS at p-value threshold (PT) of 0.05 and and WM phenotypes A) 2-back accuracy and B) 2-back dprime in the Basel 1 and Basel 2 subsample. Each result is presented twice, for PRSs including and excluding the MHC). The 0.1, 0.5 and 0.9 quantile association is presented in red, blue and green colour, respectively.

**Figure 3:**
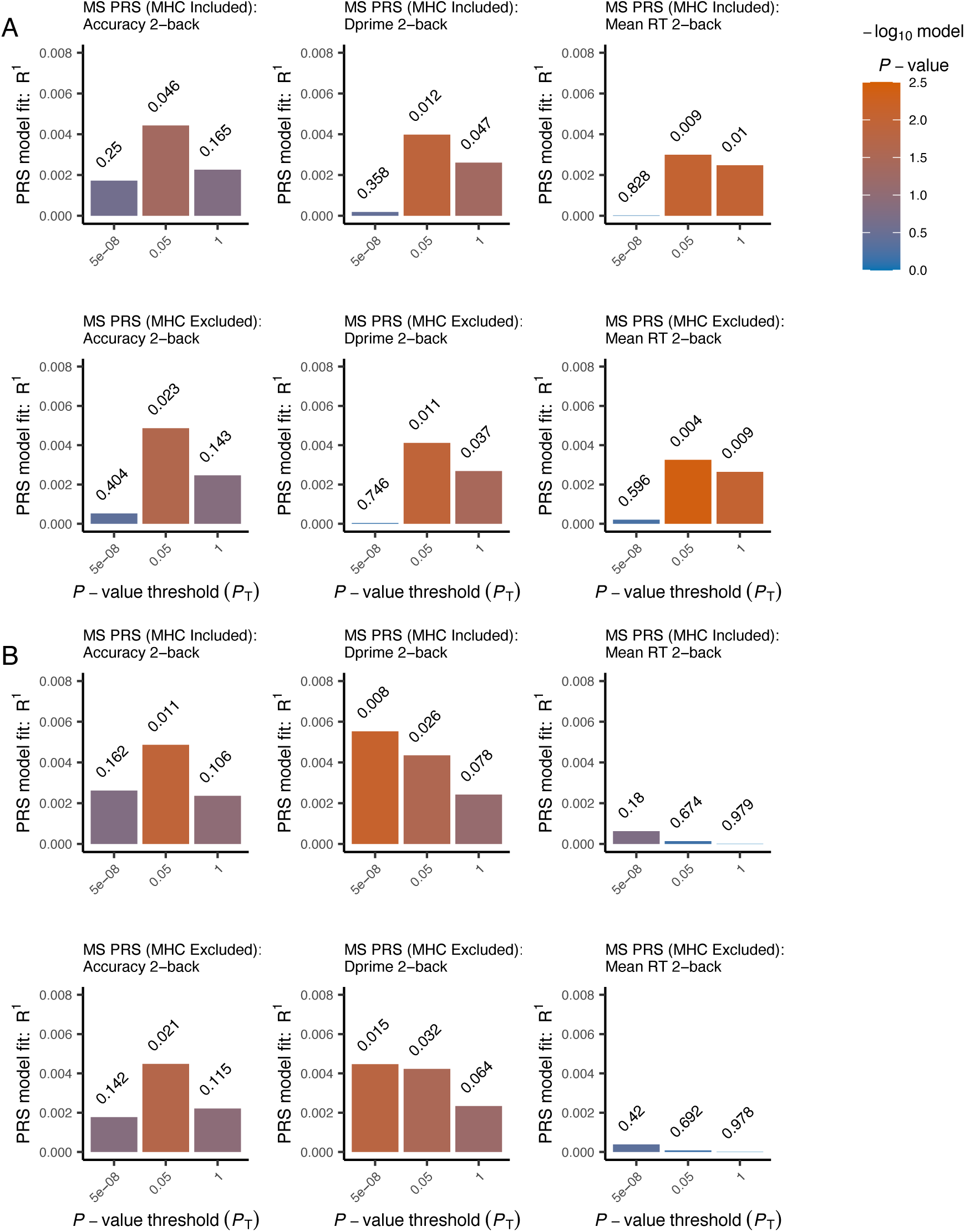
The goodness of fit (R1) of MS PRSs at different p-value thresholds (PT) for 2-back accuracy, d-prime and mean reaction time (RT), for participants within the 0.1 quantile on the respective measure in A) Basel 1 and B) Basel 2. Each result is presented twice, for PRSs including and excluding the major histocompatibility complex (MHC), respectively.

**Table 1:** Association with the MS PRS (at each PRS p-value threshold – column “PRS”) in the entire sample, the Basel 1 subsample (*N* = 1803), and the Basel 2 subsample (*N* = 1479), for 2-back accuracy, d-prime and mean RT, at each quantile, respectively. The analyses were performed with scaled values. Significant associations in the entire sample after FDR multiple-testing correction are <u>underlined</u>. *Abbreviations: qt = quantile; FDR = false discovery rate; MS = Multiple Sclerosis; PRS = Polygenic risk score; RT = Reaction Time; SE = Standard Error.

| Phenotype | qt | PRS | entire sample |  |  |  | Basel 1 |  |  | Basel 2 |  |  |
| --- | --- | --- | --- | --- | --- | --- | --- | --- | --- | --- | --- | --- |
| | | | $\beta$ | SE | $p$ | $p_{\text{FDR}}$ | $\beta$ | SE | $p$ | $\beta$ | SE | $p$ |
| accuracy | 0.1 | 1 | -0.087 | 0.041 | 0.033 | 0.18 | -0.085 | 0.061 | 0.165 | -0.089 | 0.055 | 0.106 |
| accuracy | 0.5 | 1 | -0.026 | 0.018 | 0.144 | 0.336 | -0.005 | 0.025 | 0.838 | -0.048 | 0.025 | 0.06 |
| accuracy | 0.9 | 1 | 0.005 | 0.011 | 0.678 | 0.796 | -0.013 | 0.016 | 0.413 | 0.023 | 0.016 | 0.162 |
| d-prime | 0.1 | 1 | -0.078 | 0.03 | 0.008 | 0.06 | -0.083 | 0.042 | 0.047 | -0.074 | 0.042 | 0.078 |
| d-prime | 0.5 | 1 | -0.04 | 0.021 | 0.052 | 0.2 | -0.023 | 0.028 | 0.424 | -0.06 | 0.03 | 0.047 |
| d-prime | 0.9 | 1 | 0.006 | 0.027 | 0.835 | 0.867 | -0.023 | 0.036 | 0.528 | 0.04 | 0.04 | 0.318 |
| mean RT | 0.1 | 1 | -0.026 | 0.013 | 0.049 | 0.2 | -0.045 | 0.017 | 0.01 | -0.001 | 0.02 | 0.979 |
| mean RT | 0.5 | 1 | -0.023 | 0.02 | 0.249 | 0.419 | -0.023 | 0.026 | 0.373 | -0.022 | 0.03 | 0.463 |
| mean RT | 0.9 | 1 | -0.035 | 0.04 | 0.374 | 0.532 | -0.096 | 0.053 | 0.072 | 0.04 | 0.059 | 0.499 |
| <u>accuracy</u> | <u>0.1</u> | <u>0.05</u> | <u>-0.129</u> | <u>0.04</u> | <u>0.001</u> | <u>0.018</u> | <u>-0.113</u> | <u>0.057</u> | <u>0.046</u> | <u>-0.145</u> | <u>0.057</u> | <u>0.011</u> |
| accuracy | 0.5 | 0.05 | -0.026 | 0.018 | 0.149 | 0.336 | -0.025 | 0.025 | 0.312 | -0.026 | 0.025 | 0.304 |
| accuracy | 0.9 | 0.05 | 0.006 | 0.011 | 0.557 | 0.716 | -0.009 | 0.014 | 0.549 | 0.027 | 0.017 | 0.106 |
| <u>d-prime</u> | <u>0.1</u> | <u>0.05</u> | <u>-0.087</u> | <u>0.026</u> | <u>0.001</u> | <u>0.018</u> | <u>-0.086</u> | <u>0.034</u> | <u>0.012</u> | <u>-0.087</u> | <u>0.039</u> | <u>0.026</u> |
| d-prime | 0.5 | 0.05 | -0.036 | 0.021 | 0.086 | 0.258 | -0.036 | 0.028 | 0.209 | -0.037 | 0.031 | 0.242 |
| d-prime | 0.9 | 0.05 | -0.012 | 0.025 | 0.64 | 0.786 | -0.032 | 0.035 | 0.354 | 0.01 | 0.036 | 0.776 |
| mean RT | 0.1 | 0.05 | -0.02 | 0.013 | 0.115 | 0.31 | -0.046 | 0.018 | 0.009 | 0.008 | 0.018 | 0.674 |
| mean RT | 0.5 | 0.05 | -0.024 | 0.02 | 0.232 | 0.417 | -0.039 | 0.027 | 0.139 | -0.004 | 0.031 | 0.909 |
| mean RT | 0.9 | 0.05 | -0.035 | 0.039 | 0.375 | 0.532 | -0.068 | 0.051 | 0.181 | 0.014 | 0.062 | 0.821 |
| accuracy | 0.1 | 5e-8 | -0.008 | 0.04 | 0.833 | 0.867 | 0.067 | 0.058 | 0.25 | -0.078 | 0.056 | 0.162 |
| accuracy | 0.5 | 5e-8 | 0.024 | 0.019 | 0.199 | 0.384 | 0.039 | 0.025 | 0.117 | 0.005 | 0.028 | 0.856 |
| accuracy | 0.9 | 5e-8 | 0.029 | 0.011 | 0.009 | 0.06 | 0.035 | 0.015 | 0.016 | 0.02 | 0.017 | 0.228 |
| d-prime | 0.1 | 5e-8 | -0.035 | 0.026 | 0.175 | 0.364 | 0.035 | 0.038 | 0.358 | -0.092 | 0.035 | 0.008 |
| d-prime | 0.5 | 5e-8 | 0.021 | 0.021 | 0.319 | 0.506 | 0.022 | 0.028 | 0.419 | 0.019 | 0.032 | 0.556 |
| d-prime | 0.9 | 5e-8 | 0.046 | 0.024 | 0.059 | 0.2 | 0.071 | 0.035 | 0.041 | 0.022 | 0.034 | 0.528 |
| mean RT | 0.1 | 5e-8 | 0.01 | 0.012 | 0.4 | 0.539 | -0.004 | 0.017 | 0.828 | 0.021 | 0.016 | 0.18 |
| mean RT | 0.5 | 5e-8 | 0.003 | 0.019 | 0.895 | 0.895 | 0.019 | 0.026 | 0.47 | -0.019 | 0.03 | 0.522 |
| mean RT | 0.9 | 5e-8 | -0.012 | 0.038 | 0.757 | 0.852 | -0.034 | 0.051 | 0.501 | 0.016 | 0.056 | 0.779 |

### 3.3. DTI results

We investigated the association of brain-wide mean FA with sex and age, while controlling for scanning movement, using a multiple linear model. Sex and age were significantly associated with mean FA, with higher FA in older participants and males (Age: β_scaled_ = 0.09, SE = 0.031, *p* = 0.003; Sex: β_scaled_ = -0.599, SE = 0.064, *p* < 2e-16). There was no significant association between MS PRS and brain-wide mean FA (MHC _included_: β_scaled_ = -0.027, SE = 0.031, *p* = 0.387; MHC _excluded_: β_scaled_ = -0.029, SE = 0.031, *p* = 0.34.), Figures 4.A and 4.B, or between MS PRS and mean regional FA (Figures 4.C and 4.D), after FDR correction for the number of regions tested.

**Figure 4:**
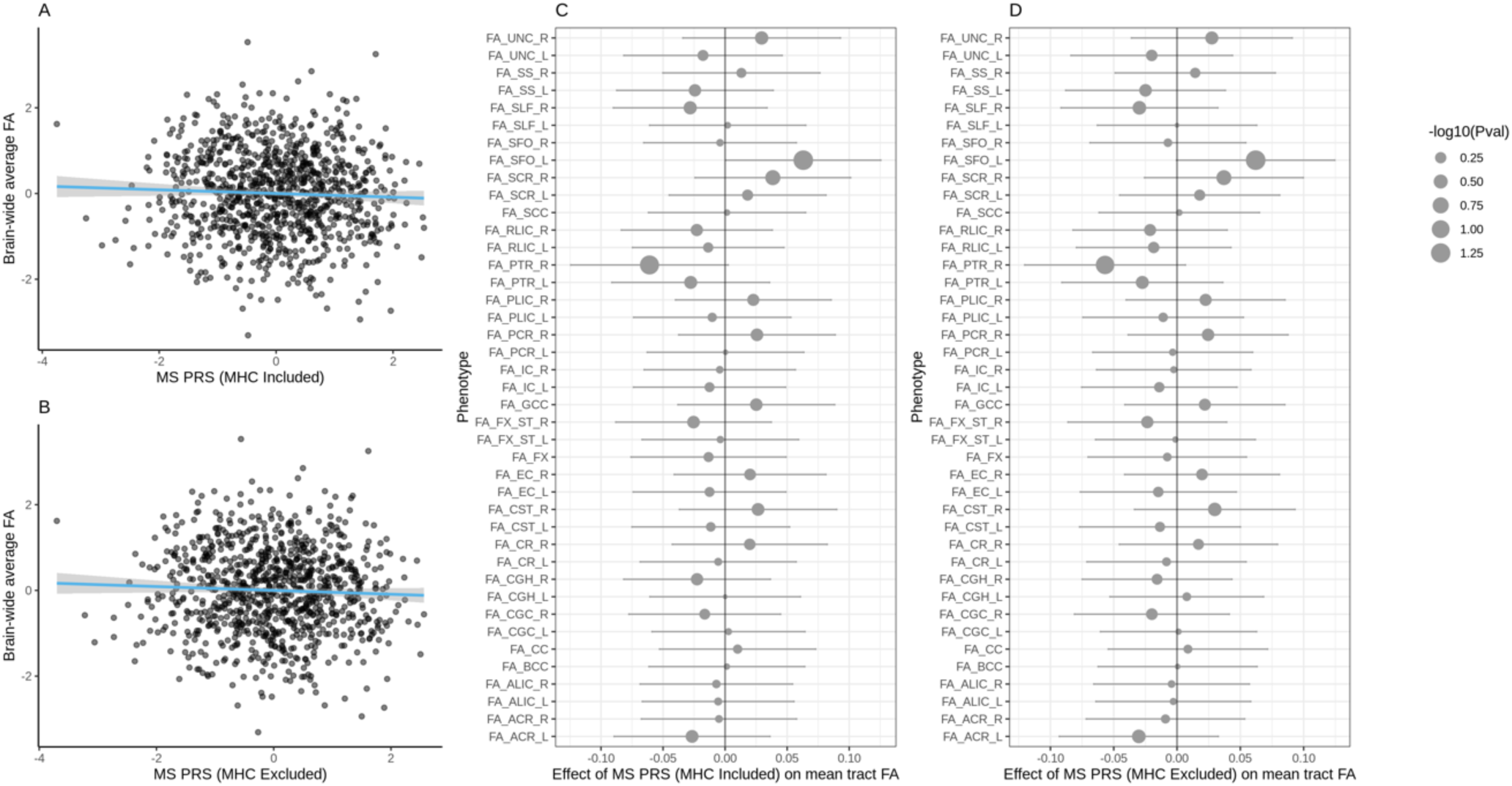
Association of MS PRS with brain-wide mean FA (3.A. – MHC included; 3.B. – MHC excluded) and MS PRS effect (beta value) on mean FA per brain tract (3.C. – MHC included; 3.D. – MHC excluded). The vertical lines in 3.C. and 3.D. represent 95 % confidence intervals. Tract name abbreviations are taken from the ENIGMA’s look-up table (Suppl. Table 2).

Brain-wide variability in number of potholes and molehills is presented in Suppl. figure 2, and closely resembled the variability reported in ^16^. We investigated the association between sex, age and the scanning movement parameter for the brain-wide number of potholes and molehills, respectively, with two multiple linear models. Participants’ sex was significantly associated with the number potholes, with fewer potholes in females, across all cluster sizes (size _25_: β_scaled_ = -0.65, SE = 0.063, *p* < 2e-16; size _50_: β_scaled_ = -0.589, SE = 0.063, *p* < 2e-16; size _100_: β_scaled_ = -0.569, SE = 0.064, *p* < 2e-16; size _200_: β_scaled_ = -0.475, SE = 0.065, *p* = 4.78e-13). There were no other significant associations for potholes or molehills (Suppl. figures 3, 4 and 5). Furthermore, there was no significant association between the MS PRS (MHC included/excluded) and the brain-wide number of potholes or molehills, (Figure 5 and Suppl. table 3), or the number of potholes or molehills per brain tract (Figure 6, Suppl. figure 6), after FDR correction for the number of brain tracts tested.

**Figure 5:**
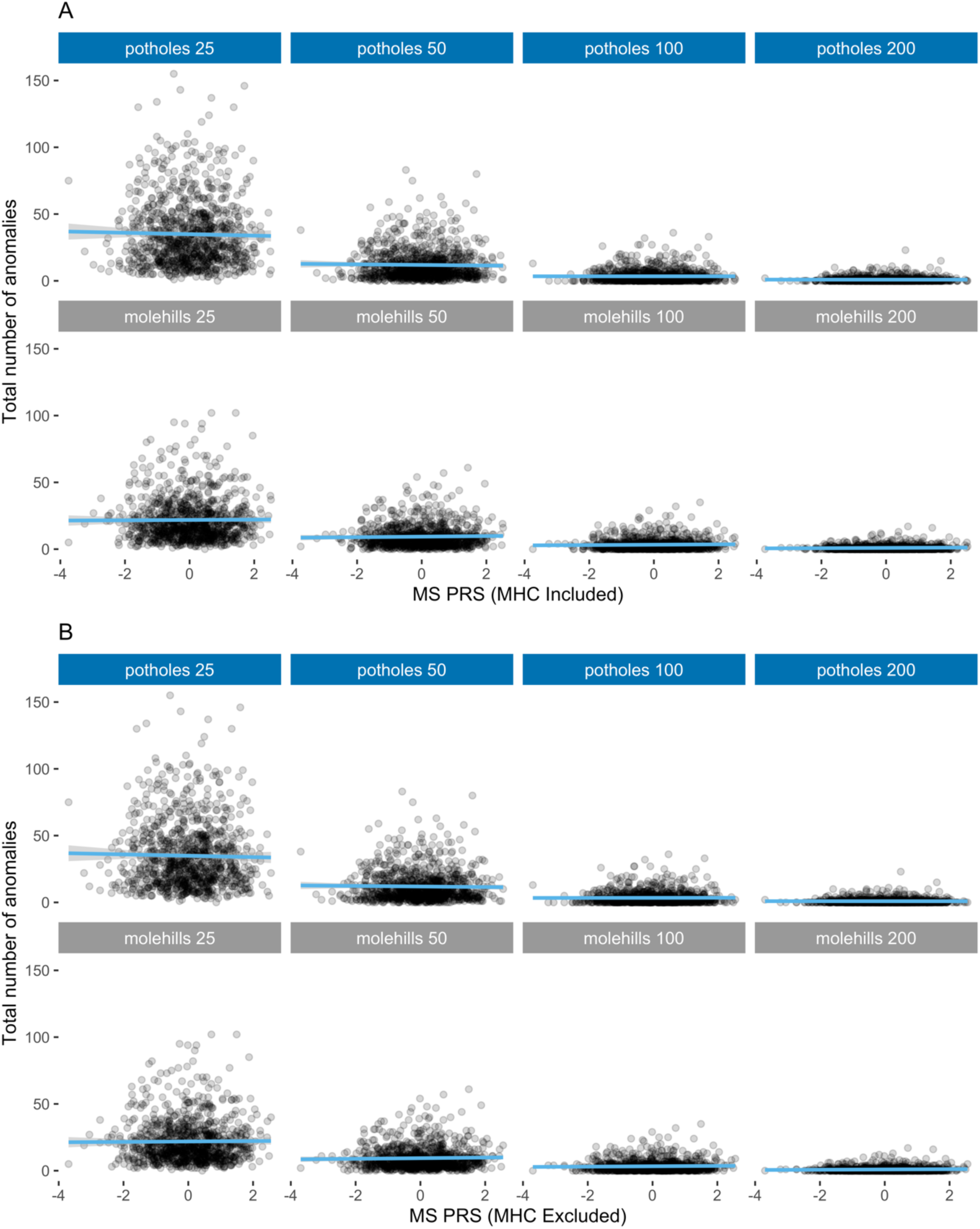
Association between the brain-wide number of potholes and molehills, per cluster size, and the MS PRS (SNP-association *p*-value threshold = 0.01) with A) MHC included B) MHC excluded.

**Figure 6:**
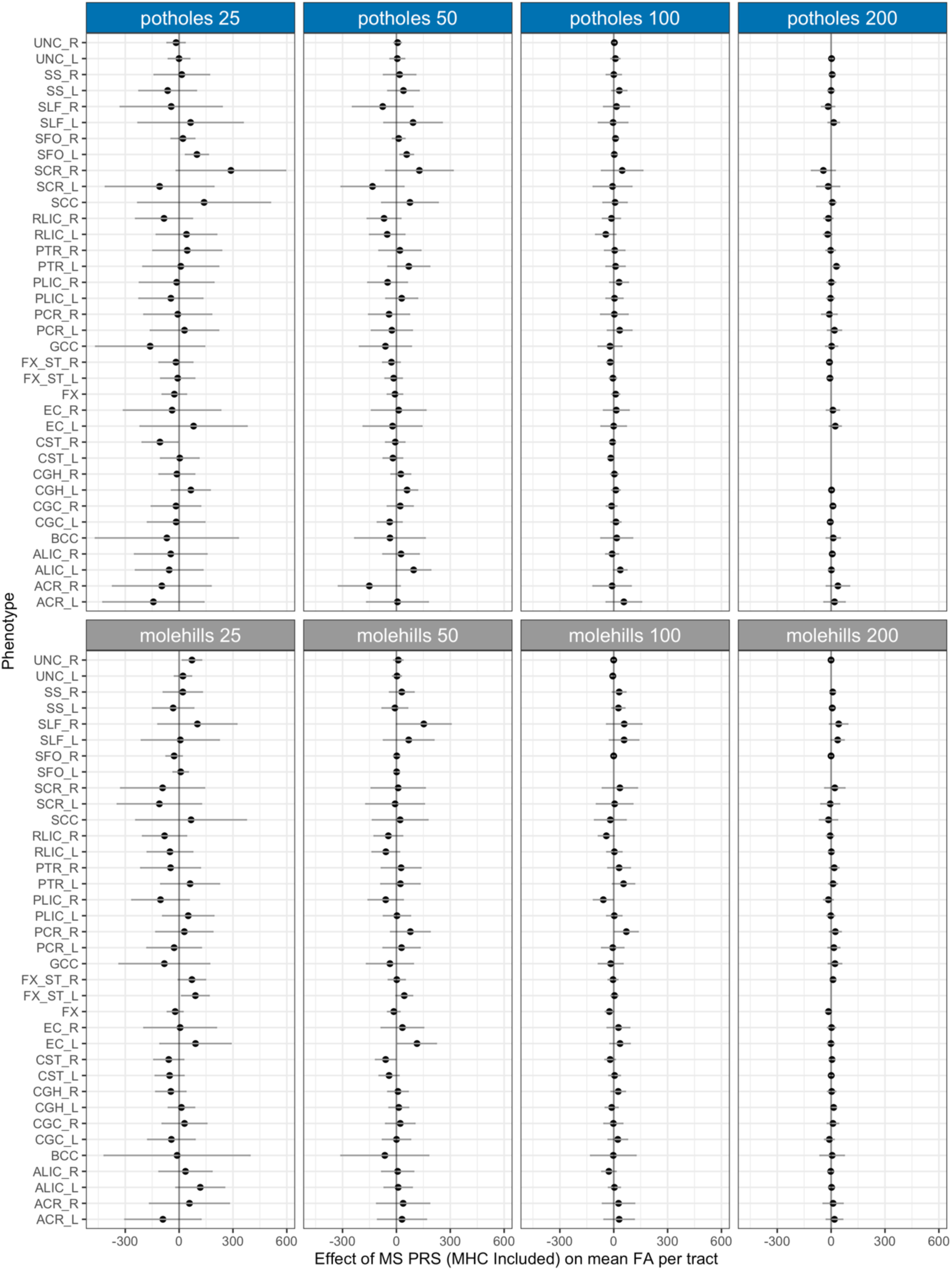
Association between the number of potholes and molehills within each brain tract, per cluster size, and the MS PRS (SNP-association *p* value threshold = 0.01), MHC included. *Note:* Missing values indicate that participants had no potholes or molehills in that tract, for the particular cluster size. Tract name abbreviations are taken from the ENIGMA’s look-up table (Suppl. Table 2). The vertical lines represent 95 % confidence intervals. For results with MHC excluded please see Suppl. Figure 6.

## 4. Discussion

In the current study, we identified a significant correlation between higher polygenic risk for MS, based on the largest GWAS to date, and lower working memory performance, especially in healthy young adults scoring withing in the lowest 10^th^ percentile on the WM task. The polygenic score including all common variants with at least nominally significant association with MS (*p* < 0.05) had the strongest WM association, indicating that variants that reached nominal significance with respect to MS risk contributed to WM performance. Importantly, the direction of effect was consistent between our two subsamples and the effect sizes were similar. The effect was nominally significant in both subsamples. Furthermore, we found no consistent MS polygenic risk association across the subsamples for 2-back mean RT, 0-back mean RT, accuracy, and d-prime, respectively, indicating no significant secondary effects on attention and processing speed.

The identified link between the polygenic MS risk and WM is interesting but remains to be replicated in future studies. Of note, while the polygenic scores calculated in this study were based on a MS GWAS meta-analysis of over 40’000 individuals, this sample size is still modest compared to GWAS data from other diseases commonly used for polygenic scoring, e.g., ^29, 30^. Associating the polygenic MS risk with WM performance may contribute to identification of molecular pathways linked to cognitive deficits in MS. While the PRS calculated in this study captures general, i.e., biologically broad genetic liability, replicating the analysis in larger samples would allow to additionally calculate pathway-based PRSs ^31^ and potentially identify distinct gene groups enriched for WM-related MS genetic burden.

There are some noteworthy limitations stemming from our sample and task characteristics. The average 2-back task performance was skewed towards high accuracy in both subsamples (Figure 1). We addressed this issue by using quantile regression, which is suitable for skewed samples ^23^, and we were able to identify a significant effect of the polygenic MS risk on WM based on the 2-back condition in the lowest 10 % of task performers. However, this effect may have been attenuated by the limitation of phenotypic variability in the behavioural measurement in the overall sample. Future studies using a more difficult task, e.g., 3-back, might better capture WM variability across individuals with higher performance levels. Additionally, it is not clear if the link between the polygenic MS risk and WM identified in the current study can already be seen in adolescence and childhood, or if it persists later in life. Moreover, all data used in this study were from participants of European ancestry. While this is the case for the gross majority of genetic studies on MS to date, there are growing efforts to extend the research in this field to different ancestral backgrounds ^32, 33^. It remains to be seen whether the here identified link between polygenic MS risk and WM can be generalized to individuals from different ancestries.

We also investigated if the effect of polygenic MS risk on brain-structural FA measurements identified in children from the general population can be generalized to healthy young adults. We did not find such an effect on average brain-wide FA, or on average FA per tract in our sample. This is in-line with recent results from a large sample of adults over 40 years of age ^17^, indicating that the effect may be limited to childhood (and possibly adolescence). Of note, using the ‘pothole’ method for identification of non-spatially overlapping white matter abnormalities, as in ^16^, we could also not find differences related to polygenic risk for MS in our adult sample. Some of the potential reasons for a discrepancy between findings in children and adults have been previously stated. For example, according to de Mol et al., (2022) ^19^ the link between the polygenic MS risk and FA in children may be due to age-related white matter maturation, no longer captured by the FA measures at the developmental endpoint. While some regional FA changes can still be observed during early adulthood ^34^, the polygenic MS risk association may be specific to an earlier developmental stage and therefore no longer detectable within the 18-35-year-old age group.

In summary, by identifying a link between the polygenic risk for MS and working memory performance in healthy, low-performing individuals, this study contributes to the understanding of the genetic architecture of MS. Whether this relationship extends to other populations and WM tasks remains to be seen. Furthermore, by not replicating the association between polygenic MS risk and FA measurements found in children, using the same analytical approach and a comparable sample size, the current study indicates that such association may be either attenuated or no longer existing in early adulthood.

## Data Availability

The multiple sclerosis GWAS data can be obtained upon request at https://imsgc.net/. Other data used in this paper are available upon reasonable request to the authors.

## 5. Acknowledgements

We would like to thank the IMSGC for kindly sharing their MS GWAS data with us for the purpose of this study.

## 8. Data availability

The MS GWAS data can be obtained upon request at https://imsgc.net/. For access to other data used in this paper, please contact the authors.

