## Supplementary material for "Polygenic susceptibility for multiple sclerosis is associated with working memory in low-performing young adults"

**Affiliations:**

### 1. DTI

#### 1.1 Acquisition

Brain imaging was performed on a Siemens Magnetom Verio 3T whole-body MR unit equipped with a twelve-channel head coil. Diffusion volumes were acquired by using a single-shot echo-planar sequence, and consisted of 64 diffusion-weighted volumes (b = 900 s/mm$^2$) and one unweighted volume (b = 0). Acquisition parameters were as follows: TR=9000 ms, TE=82 ms, FOV=320 mm, GRAPPA R=2.0, voxel size 2.5 × 2.5 × 2.5 mm$^3$. Furthermore, a high-resolution T1-weighted anatomical image was acquired with a magnetization prepared gradient echo sequence (MPRAGE, TR = 2000 ms; TE = 3.37 ms; TI = 1,000 ms; flip angle=8; 176 sagittal slices; FOV = 256 mm; voxel size 1 × 1 × 1 mm$^3$).

#### 1.2 Pre-processing

Pre-processing of the diffusion data was based on ENIGMA's DTI protocols (https://enigma.ini.usc.edu/protocols/dti-protocols/), using the FMRIB Software Library (FSL, v.6.0.2). Movement and eddy currents corrections [2] were combined with a noise reduction technique [3]. Each subject's high resolution T1 image was mapped to the diffusion's unweighted volume, in order to correct for EPI-induced distortions. Because of the inherent non-linear nature of those distortions, this mapping was computed using a non-linear approach (ANTs, v.2.1.0, [4]) using information from the unweighted volume (b0, code in Appendix 1). DTI metrics were consequently computed using the T1-warped data. The following metrics were computed [5]: fractional anisotropy (FA), mode of anisotropy (MO), mean diffusivity (MD), axial and radial diffusivity (AD and RD, respectively). FA was the main measurement of interest, and was used for subject-to-template normalization (code in Appendix 2). The template was ENIGMA-DTI's FA template (http://enigma.ini.usc.edu/wp-content/uploads/2013/02/enigmaDTI.zip). Each subject's image was then skeletonized using ENIGMA's skeleton and the tbss_skeleton command. Finally, mean DTI metrics values were extracted based on the skeletonized images. This resulted in 46 mean values per participant and metric, following regions of interest defined in the JHU DTI-based white-matter atlas (Suppl. table 2). The DTI sample consisted of 1122 participants from Basel 2.

2. Supplementary Tables

| | | | entire sample | | | | Basel 1 | | | Basel 2 | | |
|---|---|---|---|---|---|---|---|---|---|---|---|---|
| Phenotype | qt | PRS | β | SE | *p* | *p* FDR | β | SE | *p* | β | SE | *p* |
| accuracy | 0.1 | 1 | -0.088 | 0.041 | 0.031 | 0.181 | -0.084 | 0.057 | 0.143 | -0.092 | 0.058 | 0.115 |
| accuracy | 0.5 | 1 | -0.026 | 0.018 | 0.14 | 0.302 | -0.005 | 0.025 | 0.841 | -0.047 | 0.025 | 0.058 |
| accuracy | 0.9 | 1 | 0.004 | 0.011 | 0.695 | 0.816 | -0.013 | 0.016 | 0.394 | 0.022 | 0.016 | 0.159 |
| d-prime | 0.1 | 1 | -0.081 | 0.029 | 0.005 | 0.048 | -0.087 | 0.041 | 0.037 | -0.075 | 0.041 | 0.064 |
| d-prime | 0.5 | 1 | -0.041 | 0.021 | 0.046 | 0.181 | -0.023 | 0.028 | 0.425 | -0.063 | 0.03 | 0.039 |
| d-prime | 0.9 | 1 | 0.002 | 0.027 | 0.926 | 0.926 | -0.028 | 0.036 | 0.444 | 0.039 | 0.04 | 0.329 |
| mean RT | 0.1 | 1 | -0.026 | 0.013 | 0.045 | 0.181 | -0.045 | 0.017 | 0.009 | -0.001 | 0.02 | 0.978 |
| mean RT | 0.5 | 1 | -0.023 | 0.02 | 0.249 | 0.448 | -0.023 | 0.026 | 0.382 | -0.022 | 0.029 | 0.453 |
| mean RT | 0.9 | 1 | -0.035 | 0.04 | 0.381 | 0.557 | -0.096 | 0.053 | 0.074 | 0.039 | 0.059 | 0.506 |
| accuracy | 0.1 | 0.05 | -0.129 | 0.04 | 0.001 | 0.016 | -0.125 | 0.055 | 0.023 | -0.134 | 0.058 | 0.021 |
| accuracy | 0.5 | 0.05 | -0.028 | 0.018 | 0.12 | 0.302 | -0.025 | 0.025 | 0.318 | -0.031 | 0.026 | 0.229 |
| accuracy | 0.9 | 0.05 | 0.005 | 0.011 | 0.669 | 0.816 | -0.013 | 0.014 | 0.374 | 0.028 | 0.017 | 0.093 |
| d-prime | 0.1 | 0.05 | -0.085 | 0.026 | 0.001 | 0.016 | -0.085 | 0.033 | 0.011 | -0.085 | 0.04 | 0.032 |
| d-prime | 0.5 | 0.05 | -0.038 | 0.021 | 0.069 | 0.207 | -0.036 | 0.028 | 0.201 | -0.04 | 0.031 | 0.195 |
| d-prime | 0.9 | 0.05 | -0.022 | 0.025 | 0.383 | 0.557 | -0.046 | 0.034 | 0.173 | 0.008 | 0.038 | 0.838 |
| mean RT | 0.1 | 0.05 | -0.025 | 0.013 | 0.055 | 0.185 | -0.049 | 0.017 | 0.004 | 0.008 | 0.02 | 0.692 |
| mean RT | 0.5 | 0.05 | -0.026 | 0.02 | 0.191 | 0.369 | -0.038 | 0.027 | 0.152 | -0.011 | 0.03 | 0.719 |
| mean RT | 0.9 | 0.05 | -0.032 | 0.037 | 0.392 | 0.557 | -0.072 | 0.051 | 0.158 | 0.014 | 0.054 | 0.798 |
| accuracy | 0.1 | 5e-8 | -0.019 | 0.037 | 0.61 | 0.784 | 0.046 | 0.055 | 0.404 | -0.075 | 0.051 | 0.142 |
| accuracy | 0.5 | 5e-8 | 0.016 | 0.018 | 0.382 | 0.557 | 0.03 | 0.024 | 0.215 | -0.003 | 0.028 | 0.926 |
| accuracy | 0.9 | 5e-8 | 0.022 | 0.011 | 0.047 | 0.181 | 0.023 | 0.015 | 0.115 | 0.02 | 0.017 | 0.225 |
| d-prime | 0.1 | 5e-8 | -0.04 | 0.026 | 0.125 | 0.302 | 0.012 | 0.038 | 0.746 | -0.087 | 0.036 | 0.015 |
| d-prime | 0.5 | 5e-8 | 0.011 | 0.021 | 0.599 | 0.784 | 0.016 | 0.028 | 0.569 | 0.005 | 0.032 | 0.885 |
| d-prime | 0.9 | 5e-8 | 0.037 | 0.026 | 0.146 | 0.302 | 0.063 | 0.037 | 0.086 | 0.013 | 0.036 | 0.716 |
| mean RT | 0.1 | 5e-8 | 0.002 | 0.013 | 0.867 | 0.926 | -0.009 | 0.018 | 0.596 | 0.015 | 0.019 | 0.42 |
| mean RT | 0.5 | 5e-8 | 0.003 | 0.019 | 0.892 | 0.926 | 0.02 | 0.025 | 0.435 | -0.021 | 0.029 | 0.483 |
| mean RT | 0.9 | 5e-8 | -0.01 | 0.038 | 0.79 | 0.889 | -0.033 | 0.05 | 0.508 | 0.02 | 0.057 | 0.726 |

**Suppl. table 1**

Association with the MS PRS - **MHC region excluded** (at each PRS p-value threshold – column "PRS") in the entire sample, the Basel 1 subsample (*N* = 1803), and the Basel 2 subsample (*N* = 1479), for 2-back accuracy, d-prime and mean RT, at each quantile, respectively. The analyses were performed with scaled values.

*Abbreviations: qt = quantile; FDR = false discovery rate; MS = Multiple Sclerosis; PRS = Polygenic risk score; RT = Reaction Time; SE = Standard Error.

| | | | entire sample | | | | Basel 1 | | | Basel 2 | | |
|---|---|---|---|---|---|---|---|---|---|---|---|---|
| Phenotype | qt | PRS | β | SE | *p* | *p* FDR | β | SE | *p* | β | SE | *p* |
| accuracy | 0.1 | 1 | -0.017 | 0.041 | 0.675 | 0.81 | -0.092 | 0.061 | 0.127 | 0.047 | 0.056 | 0.401 |
| accuracy | 0.5 | 1 | -0.014 | 0.015 | 0.351 | 0.768 | -0.027 | 0.021 | 0.191 | 0 | 0.021 | 1 |
| d-prime | 0.1 | 1 | -0.024 | 0.036 | 0.511 | 0.768 | -0.083 | 0.051 | 0.103 | 0.036 | 0.051 | 0.482 |
| d-prime | 0.5 | 1 | -0.014 | 0.021 | 0.512 | 0.768 | -0.029 | 0.028 | 0.307 | 0.005 | 0.032 | 0.874 |
| mean RT | 0.1 | 1 | -0.017 | 0.016 | 0.273 | 0.768 | -0.018 | 0.019 | 0.34 | -0.015 | 0.028 | 0.586 |
| mean RT | 0.5 | 1 | -0.027 | 0.016 | 0.102 | 0.768 | -0.026 | 0.022 | 0.222 | -0.027 | 0.025 | 0.276 |
| accuracy | 0.1 | 0.05 | -0.009 | 0.039 | 0.821 | 0.821 | -0.052 | 0.056 | 0.349 | 0.031 | 0.054 | 0.56 |
| accuracy | 0.5 | 0.05 | -0.013 | 0.015 | 0.394 | 0.768 | -0.023 | 0.02 | 0.251 | 0 | 0.022 | 1 |
| d-prime | 0.1 | 0.05 | -0.014 | 0.033 | 0.672 | 0.81 | -0.043 | 0.049 | 0.382 | 0.01 | 0.045 | 0.817 |
| d-prime | 0.5 | 0.05 | -0.02 | 0.021 | 0.355 | 0.768 | -0.021 | 0.028 | 0.467 | -0.019 | 0.033 | 0.566 |
| mean RT | 0.1 | 0.05 | 0.012 | 0.016 | 0.448 | 0.768 | -0.003 | 0.02 | 0.871 | 0.043 | 0.028 | 0.122 |
| mean RT | 0.5 | 0.05 | -0.008 | 0.016 | 0.637 | 0.81 | -0.031 | 0.022 | 0.163 | 0.021 | 0.025 | 0.39 |
| accuracy | 0.1 | 5e-8 | -0.011 | 0.036 | 0.768 | 0.813 | -0.025 | 0.052 | 0.633 | 0.003 | 0.05 | 0.957 |
| accuracy | 0.5 | 5e-8 | 0.005 | 0.015 | 0.753 | 0.813 | 0.009 | 0.021 | 0.665 | 0 | 0.022 | 1 |
| d-prime | 0.1 | 5e-8 | -0.029 | 0.034 | 0.386 | 0.768 | -0.054 | 0.045 | 0.231 | 0.002 | 0.051 | 0.969 |
| d-prime | 0.5 | 5e-8 | 0.018 | 0.021 | 0.4 | 0.768 | 0.012 | 0.028 | 0.663 | 0.024 | 0.031 | 0.44 |
| mean RT | 0.1 | 5e-8 | 0.028 | 0.014 | 0.056 | 0.768 | 0.006 | 0.019 | 0.748 | 0.059 | 0.023 | 0.01 |
| mean RT | 0.5 | 5e-8 | 0.018 | 0.016 | 0.268 | 0.768 | -0.001 | 0.022 | 0.966 | 0.041 | 0.024 | 0.091 |

**Suppl. table 2**

Association with the MS PRS - **MHC region included** (at each PRS p-value threshold – column "PRS") in the entire sample, the Basel 1 subsample ($N$ = 1803), and the Basel 2 subsample ($N$ = 1479), for **0-back** accuracy, d-prime and mean RT, at the 0.1 and the 0.5 quantile, respectively. Associations were not calculated at the 0.9 quantile, due to a lack of phenotypic variability. The analyses were performed with scaled values.

*Abbreviations: qt = quantile; FDR = false discovery rate; MS = Multiple Sclerosis; PRS = Polygenic risk score; RT = Reaction Time; SE = Standard Error.

| | | | entire sample | | | | Basel 1 | | | Basel 2 | | |
|---|---|---|---|---|---|---|---|---|---|---|---|---|
| Phenotype | qt | PRS | β | SE | *p* | *p* FDR | β | SE | *p* | β | SE | *p* |
| accuracy | 0.1 | 1 | -0.005 | 0.042 | 0.909 | 0.963 | -0.095 | 0.06 | 0.114 | 0.079 | 0.058 | 0.172 |
| accuracy | 0.5 | 1 | -0.014 | 0.015 | 0.331 | 0.825 | -0.028 | 0.021 | 0.172 | 0 | 0.021 | 1 |
| d-prime | 0.1 | 1 | -0.02 | 0.035 | 0.562 | 0.825 | -0.088 | 0.052 | 0.087 | 0.036 | 0.047 | 0.442 |
| d-prime | 0.5 | 1 | -0.015 | 0.021 | 0.474 | 0.825 | -0.031 | 0.029 | 0.277 | 0.004 | 0.032 | 0.892 |
| mean RT | 0.1 | 1 | -0.016 | 0.015 | 0.298 | 0.825 | -0.02 | 0.018 | 0.281 | -0.007 | 0.027 | 0.792 |
| mean RT | 0.5 | 1 | -0.028 | 0.016 | 0.08 | 0.825 | -0.027 | 0.022 | 0.215 | -0.031 | 0.025 | 0.214 |
| accuracy | 0.1 | 0.05 | -0.025 | 0.038 | 0.506 | 0.825 | -0.082 | 0.054 | 0.126 | 0.033 | 0.055 | 0.542 |
| accuracy | 0.5 | 0.05 | -0.014 | 0.015 | 0.35 | 0.825 | -0.025 | 0.02 | 0.202 | 0 | 0.021 | 1 |
| d-prime | 0.1 | 0.05 | -0.015 | 0.033 | 0.642 | 0.825 | -0.046 | 0.049 | 0.346 | 0.011 | 0.045 | 0.81 |
| d-prime | 0.5 | 0.05 | -0.022 | 0.021 | 0.309 | 0.825 | -0.023 | 0.028 | 0.408 | -0.019 | 0.032 | 0.55 |
| mean RT | 0.1 | 0.05 | 0.008 | 0.016 | 0.627 | 0.825 | -0.01 | 0.019 | 0.614 | 0.044 | 0.028 | 0.114 |
| mean RT | 0.5 | 0.05 | -0.01 | 0.017 | 0.568 | 0.825 | -0.031 | 0.023 | 0.171 | 0.016 | 0.025 | 0.508 |
| accuracy | 0.1 | 5e-8 | 0 | 0.036 | 0.996 | 0.996 | -0.058 | 0.05 | 0.248 | 0.059 | 0.051 | 0.244 |
| accuracy | 0.5 | 5e-8 | -0.003 | 0.016 | 0.857 | 0.963 | -0.005 | 0.022 | 0.802 | 0 | 0.022 | 1 |
| d-prime | 0.1 | 5e-8 | -0.021 | 0.032 | 0.513 | 0.825 | -0.065 | 0.047 | 0.163 | 0.017 | 0.043 | 0.689 |
| d-prime | 0.5 | 5e-8 | 0.006 | 0.021 | 0.778 | 0.933 | -0.006 | 0.029 | 0.845 | 0.02 | 0.032 | 0.525 |
| mean RT | 0.1 | 5e-8 | 0.021 | 0.015 | 0.147 | 0.825 | 0.005 | 0.018 | 0.776 | 0.051 | 0.025 | 0.039 |
| mean RT | 0.5 | 5e-8 | 0.008 | 0.017 | 0.621 | 0.825 | -0.001 | 0.022 | 0.967 | 0.021 | 0.026 | 0.415 |

**Suppl. table 3**

Association with the MS PRS - **MHC region excluded** (at each PRS p-value threshold – column "PRS") in the entire sample, the Basel 1 subsample (*N* = 1803), and the Basel 2 subsample (*N* = 1479), for **0-back** accuracy, d-prime and mean RT, at the 0.1 and the 0.5 quantile, respectively. Associations were not calculated at the 0.9 quantile, due to a lack of phenotypic variability. The analyses were performed with scaled values.

*Abbreviations: qt = quantile; FDR = false discovery rate; MS = Multiple Sclerosis; PRS = Polygenic risk score; RT = Reaction Time; SE = Standard Error.

| ROI | Abbreviation | Name |
|---|---|---|
| 1 | MCP | Middle cerebellar peduncle |
| 2 | PCT | Pontine crossing tract (a part of MCP) |
| 3 | GCC | Genu of corpus callosum |
| 4 | BCC | Body of corpus callosum |
| 5 | SCC | Splenium of corpus callosum |
| 6 | FX | Fornix (column and body of fornix) |
| 7 | CST_R | Corticospinal tract right |
| 8 | CST_L | Corticospinal tract left |
| 9 | ML_R | Medial lemniscus right |
| 10 | ML_L | Medial lemniscus left |
| 11 | ICP_R | Inferior cerebellar peduncle right |
| 12 | ICP_L | Inferior cerebellar peduncle left |
| 13 | SCP_R | Superior cerebellar peduncle right |
| 14 | SCP_L | Superior cerebellar peduncle left |
| 15 | CP_R | Cerebral peduncle right |
| 16 | CP_L | Cerebral peduncle left |
| 17 | ALIC_R | Anterior limb of internal capsule right |
| 18 | ALIC_L | Anterior limb of internal capsule left |
| 19 | PLIC_R | Posterior limb of internal capsule right |
| 20 | PLIC_L | Posterior limb of internal capsule left |
| 21 | RLIC_R | Retrolenticular part of internal capsule right |
| 22 | RLIC_L | Retrolenticular part of internal capsule left |
| 23 | ACR_R | Anterior corona radiata right |
| 24 | ACR_L | Anterior corona radiata left |
| 25 | SCR_R | Superior corona radiata right |
| 26 | SCR_L | Superior corona radiata left |
| 27 | PCR_R | Posterior corona radiata right |
| 28 | PCR_L | Posterior corona radiata left |
| 29 | PTR_R | Posterior thalamic radiation (include optic radiation) right |
| 30 | PTR_L | Posterior thalamic radiation (include optic radiation) left |
| 31 | SS_R | Sagittal stratum (include inferior longitudinal fasciculus and inferior fronto_occipital fasciculus) right |
| 32 | SS_L | Sagittal stratum (include inferior longitudinal fasciculus and inferior fronto_occipital fasciculus) left |
| 33 | EC_R | External capsule right |
| 34 | EC_L | External capsule left |
| 35 | CGC_R | Cingulum (cingulate gyrus) right |
| 36 | CGC_L | Cingulum (cingulate gyrus) left |
| 37 | CGH_R | Cingulum (hippocampus) right |
| 38 | CGH_L | Cingulum (hippocampus) left |
| 39 | FX_ST_R | Fornix (cres) _ Stria terminalis (cannot be resolved with current resolution) right |
| 40 | FX_ST_L | Fornix (cres) _ Stria terminalis (cannot be resolved with current resolution) left |

| | | |
|---|---|---|
| 41 | SLF_R | Superior longitudinal fasciculus right |
| 42 | SLF_L | Superior longitudinal fasciculus left |
| 43 | SFO_R | Superior fronto_occipital fasciculus (could be a part of anterior internal capsule) right |
| 44 | SFO_L | Superior fronto_occipital fasciculus (could be a part of anterior internal capsule) left |
| 45 | IFO_R | Inferior fronto_occipital fasciculus right |
| 46 | IFO_L | Inferior fronto_occipital fasciculus left |
| 47 | UNC_R | Uncinate fasciculus right |
| 48 | UNC_L | Uncinate fasciculus left |

#### Suppl. table 4

A list of full names and abbreviations of brain tracts, i.e., regions of interest (ROIs), as defined by ENIGMA. Regions number 1 and 2 were not considered by the ENIGMA consortium and were consequently excluded from our analyses.

| DTI FA phenotype (Cluster size) | MHC included | | | MHC excluded | | |
|---|---|---|---|---|---|---|
| | β | SE | *p* | β | SE | *p* |
| potholes (25) | -0.001 | 0.03 | 0.967 | -0.001 | 0.03 | 0.967 |
| potholes (50) | 0.002 | 0.031 | 0.942 | 0.003 | 0.031 | 0.929 |
| potholes (100) | 0.021 | 0.031 | 0.499 | 0.024 | 0.031 | 0.424 |
| potholes (200) | 0.015 | 0.031 | 0.621 | 0.014 | 0.031 | 0.641 |
| molehills (25) | 0.004 | 0.032 | 0.911 | 0.005 | 0.032 | 0.882 |
| molehills (50) | 0.02 | 0.032 | 0.532 | 0.022 | 0.032 | 0.499 |
| molehills (100) | 0.026 | 0.032 | 0.415 | 0.027 | 0.032 | 0.402 |
| molehills (200) | 0.051 | 0.032 | 0.117 | 0.052 | 0.032 | 0.105 |

#### Suppl. table 5

Association with the MS PRS (p-value threshold of 0.05), with and without the MHC region, for the brain-wide number of potholes and molehills, per cluster size, respectively. The analyses were performed with scaled values.

*Abbreviations: SE = Standard Error; DTI = Diffusion Tensor Imaging; FA = Fractional Anisotropy; MHC = Major Histocompatibility Complex; MS = Multiple Sclerosis; PRS = Polygenic risk score.

3. Supplementary Figures

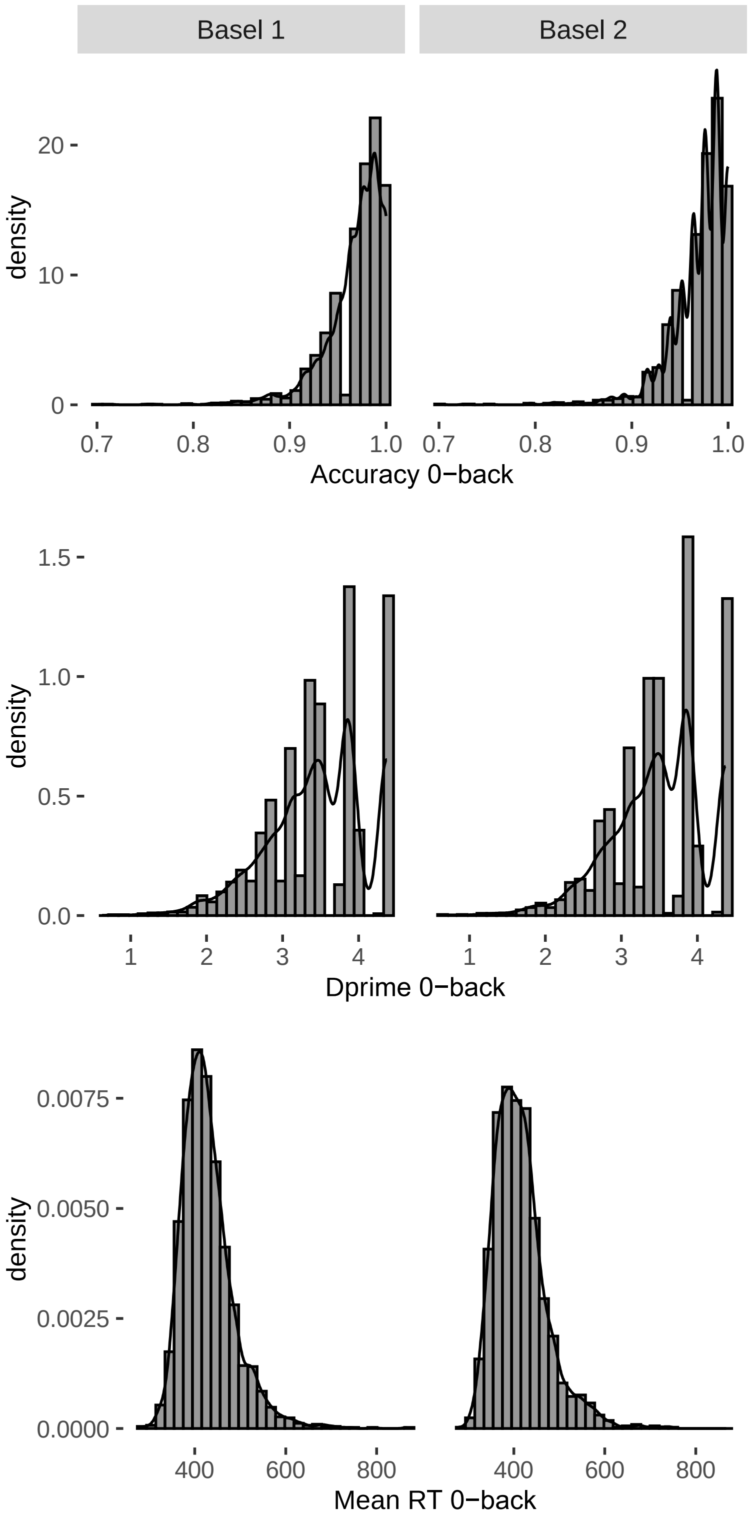

**Suppl. figure 1**

Distribution of participants' scores for accuracy, d prime and mean reaction time (RT) from the 0-back condition of the n-back task, per sample.

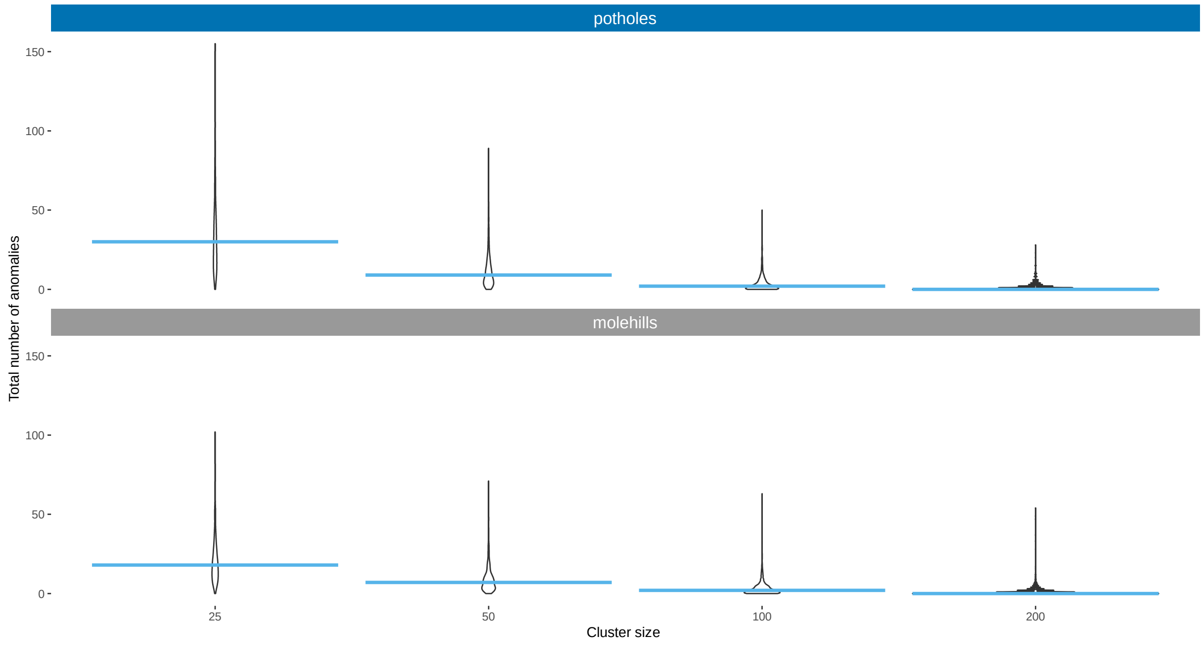

#### Suppl. figure 2

Brain-wide variability in the number of potholes and molehills, per cluster size. The straight horizontal lines represent median values.

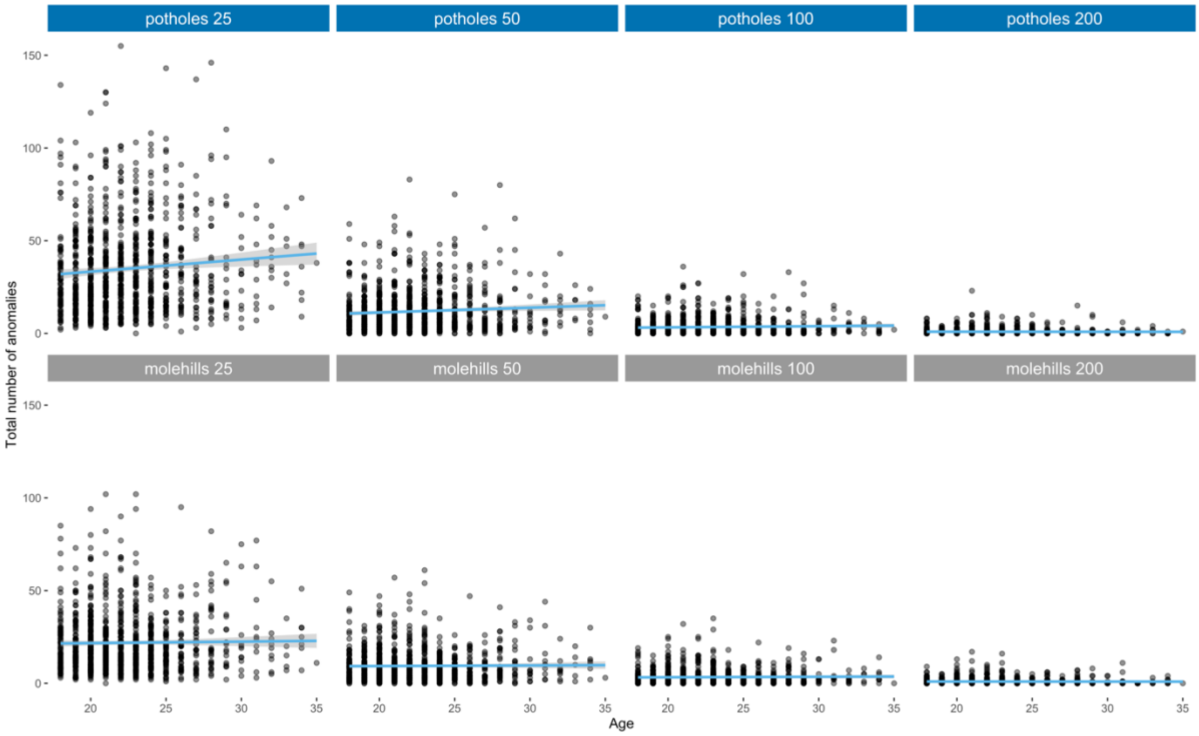

#### Suppl. figure 3

Age association for the brain-wide number of potholes and molehills, per cluster size.

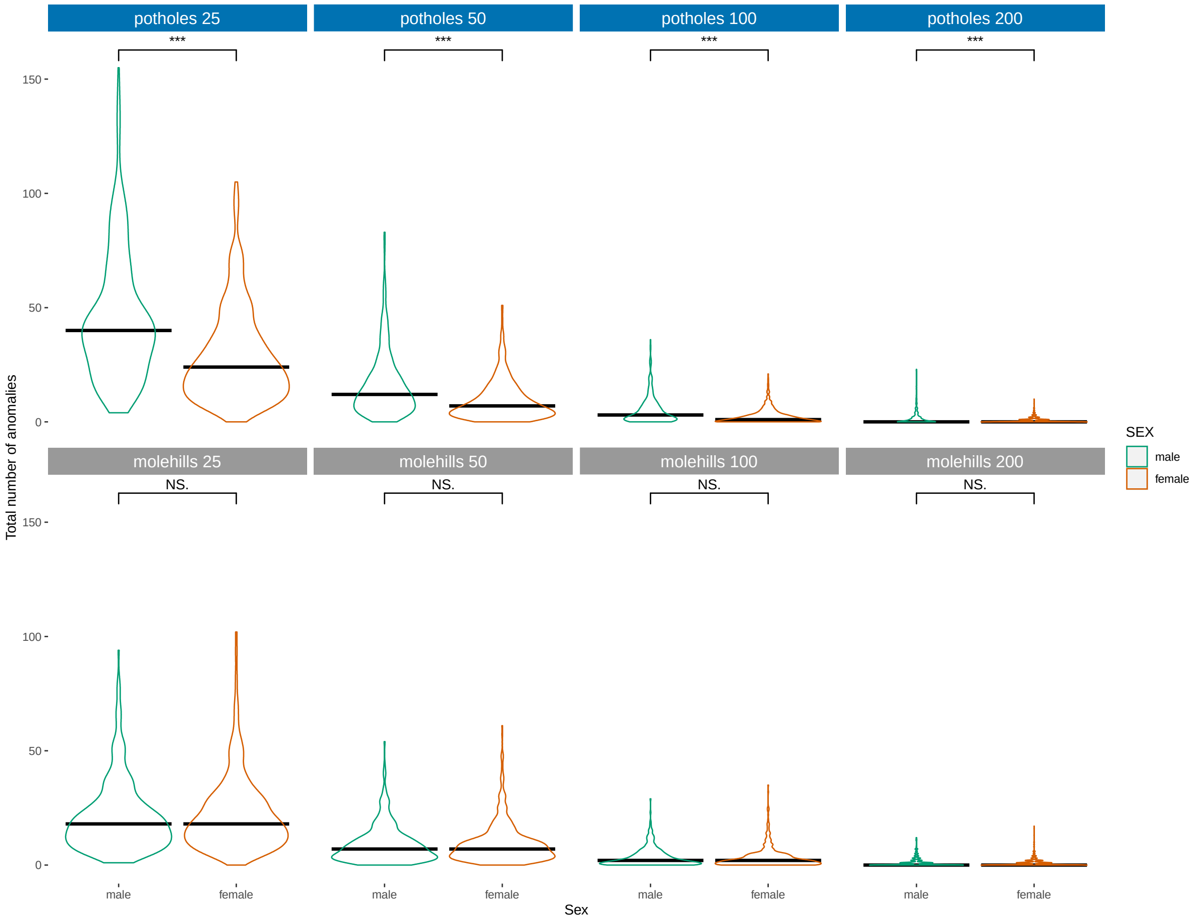

#### Suppl. figure 4

Sex association for the brain-wide number of potholes and molehills, per cluster size. The straight horizontal lines represent median values.

*Abbreviations and symbols: NS. – not significant; *** $p$-value < 0.0005

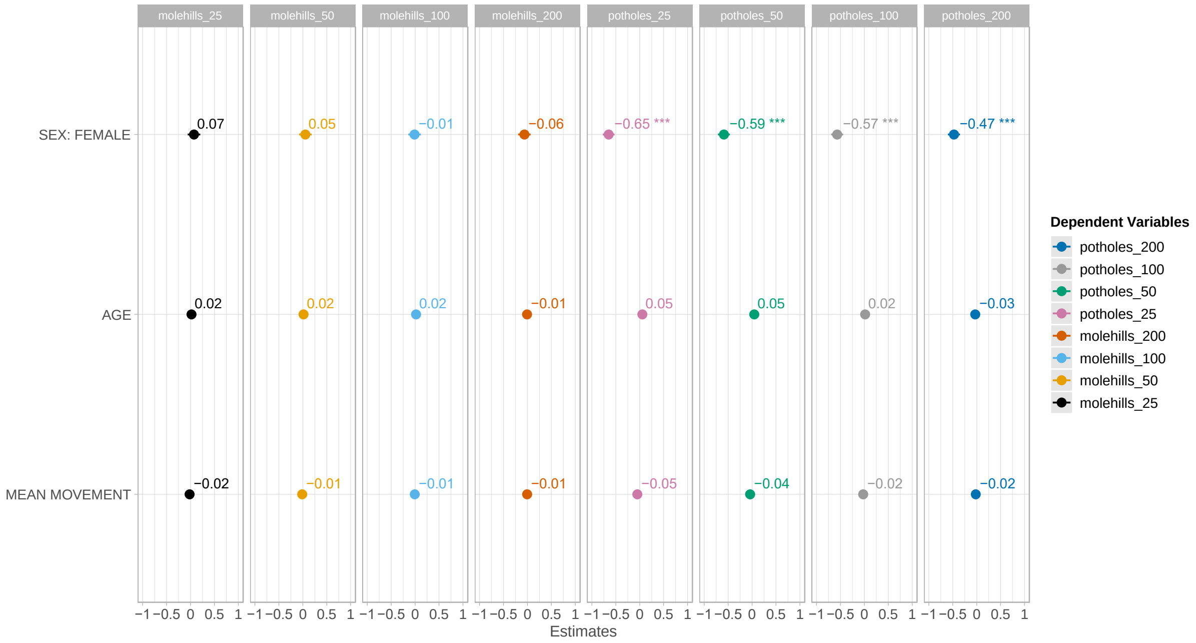

#### Suppl. figure 5

Standardized beta values of the sex, age and the scanning movement parameter association for the brain-wide number of potholes and molehills, respectively.

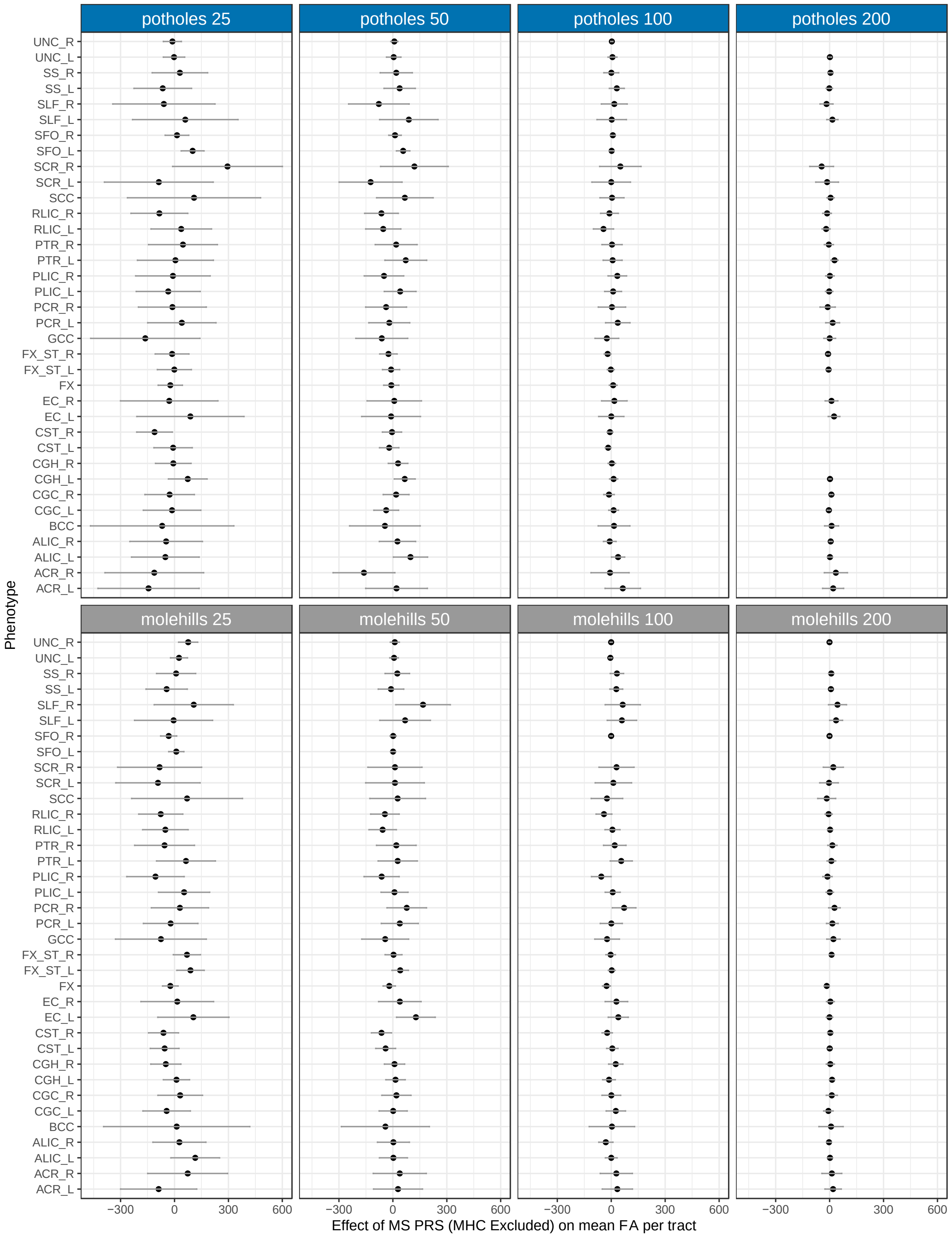

#### Suppl. figure 6

Association between the number of potholes and molehills within each brain tract, per cluster size, and the MS PRS (*p* value threshold = 0.01), MHC excluded.
Missing values indicate that participants had no potholes or molehills in that tract, for the particular cluster size. Tract name abbreviations are taken from the ENIGMA's look-up table (Suppl. table 2). The vertical lines represent 95 % confidence intervals.

### 5. Appendices

#### Appendix 1. DTI-to-T1-registration

```
# brain0N4_crop: brain-segmented T1 image
# b0_1mm: unweighted diffusion image (b=0)
antsRegistration \
                  -d 3 \
    --output b0_to_t1 \
    --metric MI[brain0N4_crop.nii.gz,b0_1mm.nii.gz, 0.5, 24, 'Regular',1] \
    --transform Affine[0.1] \
    --convergence [5x1x1,1e-6,3] \
    --shrink-factors 2x1x1 \
    --smoothing-sigmas 4x2x1 \
    --metric CC[brain0N4_crop.nii.gz,b0_1mm.nii.gz, 0.5, 4, 'Regular',1] \
    --metric MI[brain0N4_crop.nii.gz,b0_1mm.nii.gz, 0.5, 24, 'Regular',1] \
    --transform 'SyN[0.1,3.0,1.0]' \
    --convergence [5x1x1,1e-6,3] \
    --shrink-factors 2x2x1 \
    --smoothing-sigmas 3x2x1 \
    --use-histogram-matching 1 \
    --metric MI[brain0N4_crop.nii.gz,b0_1mm.nii.gz, 1, 32, 'Regular',1] \
    --transform 'SyN[0.1,3.0,0.2]' \
    --convergence [5x15x5,1e-6,3] \
    --shrink-factors 2x2x1 \
    --smoothing-sigmas 2x1x0 \
    --use-histogram-matching 1
```

#### Appendix 2. DTI subject-to-template registration

```
template="ENIGMA_DTI_FA.nii.gz"
antsRegistration \
                  -d 3 \
                  --output fa_to_template \
                  --write-composite-transform 0 \
                  -m MI[$6, fa_Warped.nii.gz, 1,24] \
                  --transform Affine[0.1] \
                  --convergence [20x20x5,1e-6,3] \
                  --shrink-factors 4x2x1 \
                  --smoothing-sigmas 4x2x1vox \
                  -m MI[$6, fa_Warped.nii.gz, 1,28] \
                  --transform SyN[0.25,3.0,0.0] \
                  --convergence [100x50x5,1e-6,5] \
                  --shrink-factors 3x2x1 \
                  --smoothing-sigmas 1.5x1x0 \
                  --use-histogram-matching 1

# combine transformations
for metric in fa md mo ad rd
do
    antsApplyTransforms \
                                    -e 3 \
        -i $4.nii.gz \
        -r $6 \
                             -t fa_to_template1Warp.nii.gz \
                                    fa_to_template0GenericAffine.mat \
                                    b0_to_t11Warp.nii.gz \
                                    b0_to_t10GenericAffine.mat \
                                    -n BSpline \
                                    -o $4_Warped.nii.gz
done
```
